# An evaluation of metabolic, dietetic, and nutritional status reveals impaired nutritional outcomes in breast cancer patients undergoing chemotherapy compared with a matched control group

**DOI:** 10.1101/2022.02.03.22270381

**Authors:** Bruna Ramos da Silva, Sarah Rufato, Mirele S. Mialich, Loris P. Cruz, Thais Gozzo, Alceu A. Jordao

## Abstract

**Purpose:** Nutritional status changes in breast cancer patients during treatment are prevalent. However, the metabolic implications of those alterations are poorly understood. We aimed to characterize body composition, lipids, glucose levels, and indices that express cardiovascular risk in breast cancer patients after completion of chemotherapy and then to compare those results with a matched control group.

**Methods:** A cross-sectional study was performed. Women who completed their chemotherapy were recruited (BC group) and compared with a group of non-malignant age- and body mass index-matched (MC group), as well as a group of healthy, non-malignant women (HC group). Body composition by bioelectrical impedance analysis, handgrip strength, and blood sample were collected. Visceral adiposity, triglyceride glucose and lipid accumulation product indices were calculated. Food consumption was assessed.

**Results:** 88 women were included (BC=36, MC=36, HC=16). BC patients demonstrated worse values of phase angle, nutritional risk index, extracellular body water to total body water ratio and lower handgrip strength. Additionally, those women had impairments in lipids, worst glucose levels, visceral fat dysfunction and consequently higher cardiovascular risk, presenting important unhealthy dietary patterns with higher carbohydrate and caloric intake and insufficient protein and fiber ingestion. No differences were observed between MC and HC.

**Conclusion:** Breast cancer patients present unhealthy metabolic, nutritional, and dietetic features when compared to a group of age- and BMI-matched non-malignant females. Also, breast cancer patients had higher levels of cardiovascular risk. Further investigations are required to examine the underlying mechanisms and the potential longitudinal changes during surveillance time.

## Introduction

Breast cancer is the most diagnosed cancer across the world, with more than 2.2 million cases in 2020 ^1^. Likewise, in Brazil breast cancer was one of the most diagnosed cancers in 2020 with 66.280 new cases ^2^. Although breast cancer is the main prevalent form of cancer, it has one of the best survival rates as well. In Brazil, the relative survival rate of 5 years between 2005 to 2009 was 87% ^3^, whereas high-income countries presented 85% to 90% during 2010 through 2014 ^1^. Considering the risk factors, this tumour is strongly associated with obesity and unhealthy body composition at the diagnosis ^4^, however, weight gain and fat mass increase can be enhanced after treatment ^5^.

Not only adiposity factors are subject to alterations by cancer treatment, but several other nutritional indicators, as lean mass, and sarcopenia ^6^, and functional capacity measured by Hand Grip Strength (HCS) ^7^ can also be affected. Phase angle (PhA), obtained by bioelectrical impedances, is considered both a prognosis and survival marker ^8–10^ and might be related to inflammatory and oxidative impairments ^11^. Its alteration has already been demonstrated as linked to an increased nutritional risk measured by the Nutritional risk index (NRI) after cancer treatment in breast cancer patients ^12^.

Similarly, metabolic changes are another possible consequence for those patients, such as lipids and glucose levels increase ^13,14^. Moreover, Godinho-Mota et al (2020) reported a visceral fat dysfunction among breast cancer patients after chemotherapy ^15^. Considering visceral fat accumulation, the adiposity indices as the visceral adiposity index (VAI), lipid accumulation product index (LAP), and triglyceride glucose index (TyG), could be important metabolic alterations tracking tools ^16–20^. Furthermore, in a previous study, our research group found a metabolic syndrome prevalence of more than 50% of breast cancer survivors ^21^, and the combination of those alterations with unhealthy body composition and improperly food intake might lead to the development of secondary illness, for instance, the cardiovascular diseases in breast cancer survivors ^22–24^, reflecting directly to the survival rates and clinical evolution after the cancer care. However, worsening in blood pressure and metabolism components are commonly identified in the association with other conditions besides cancer and the treatment, such as age, body mass index (BMI), dietetics imbalance ^25^. In particular, obesity role plays as a trigger for these alterations, in which widespread obesity is related to the increasing metabolic syndrome (MetS) cases ^26^.

Accordingly, a comparison group is important in order to identify whether the bad outcomes are associated with breast cancer and the treatment, or it is associated with age, BMI, and body fat mass amount, once those characteristics are also associated with breast cancer incidence ^27^. We hypothesized that breast cancer patients would demonstrate impairments in lipids, glucose, and body composition, which would be worse in patients compared to a matched control group of non-malignancy history females. We further hypothesized that these impairments may be explained by the presence of unhealthy body composition and dietetic inadequacy. In order to contribute to this field of knowledge, this study aims comprehensively characterize metabolism components in breast cancer patients post-chemotherapy, and to compare body composition, metabolic profile, and food intake results to non-malignant females of similar age and BMI. We also aimed to compare breast cancer patients and matched control females to a reference group of nonmalignant, healthy normal BMIs females.

## Methods

### Study Population

A cross-sectional study was performed. This study involved 88 participants: 36 patients diagnosed with early breast cancer after 1 month of chemotherapy completion (BC females), 36 non-malignant females of similar age and BMI (MC females) and 16 as a reference group of nonmalignant, healthy females (HC females) with normal BMIs (normal range). The data collection for BC was made 1 month after finalizing the chemotherapy and before the hormone therapy started, due to the possible association between hormone therapy and the increase of metabolic alterations ^28^. None of the participants have received nutritional counselling. Breast cancer patients were recruited through clinical oncology practices at Mastology ambulatory of General Hospital of School of Medicine of Ribeirão Preto, São Paulo, Brazil. During the clinical consultation, a responsible nurse informed the patient about the study. Those who were interested in knowing more about it were forwarded to talk to the study researcher. Women who met the following inclusion criteria were enrolled in the study: age ≥18 years and <65 years; a histological confirmed diagnosis of early breast cancer (range of stage I – III); completion of the breast cancer chemotherapy treatment course. Patients who previously have already received or started chemotherapy in any other moment of life; with any type of diabetes (type 1, type II or had diabetes gestational); those fitted with a defibrillator, cardiac pacemaker, metal implants or those with a local infection/wound preventing the use of bioelectric impedance analysis pads, those unable to use a handheld dynamometer due to a neuromuscular disorder were all excluded. It was adopted the breast cancer patient data collection with 1 month after finalized the chemotherapy due to the possible association between hormone therapy and the increase of MetS risk ^28^. Thus, to study only the effect of chemotherapy on the sample, the evaluation was made before the hormone therapy starting to avoid possible bias.

Women in both control groups (MC and HC) were recruited at the same hospital, the participants were employees. For both groups (BC and CG) potential participants were weighed and measured to determine BMI and completed a Health Status Screening Form to determine if they had any prior cancer or were under hormone or any other medication which could modify the metabolism that would have excluded them from participating in the study. Table 1 shows all the inclusion and exclusion criteria among the groups.

**Table 1:** Eligibility criteria for all participant groups.

| Criteria | Breast Cancer Patients | Matched Control | Health Control |
| --- | --- | --- | --- |
| **Inclusion Criteria** |  |  |  |
| **Age** | > 18 years old | Within ± 3 years of matched patient | > 18 years old <60 years old |
| **BMI** | | Within ± 2kg/m<sup>2</sup> of matched patient | 18.5 - 24.9 |
| **Sex** | female | female | female |
| **Clinical characteristics:** |  |  |  |
| *Cancer diagnosis* | Recent diagnosis of breast cancer without previous chemotherapy | No history of cancer | No history of cancer |
| *cancer stage* | Clinical stages I- III |  |  |
| *Treatment* | After completion of chemotherapy OR finished chemotherapy course |  |  |
| **Exclusion criteria** |  |  |  |
| *Metastasis* |  |  |  |
| *Previous diagnosis of cancer* |  |  |  |
| *Diabetes any type* |  |  |  |
| *HIV* |  |  |  |
| *thyroid disease that is not currently managed with medication* |  |  |  |
| *Pregnancy* |  |  |  |
| *BIA exclusion factors* |  |  |  |
| *Uncontrolled BP* |  |  |  |

Captions: BMI: Body mass index; BP: Blood Pressure.

After screening, the participants eligible for the study were scheduled for the data collection visit. The Institutional Review Board at the University of São Paulo, General Hospital, approved the current study, protocol number: HCRP 14608/2017.

### Data Collection

All participants underwent anthropometric assessments, bioelectrical impedance analysis, handgrip strength test, food intake; and blood chemical analyzes were collected. Socioeconomic, demographic, behavioral, clinical, and therapeutic data were collected directly from participants using questionnaires or obtained from medical records in the BC group. In addition, written informed consent was obtained at the begging of the visit. After 3 weeks of the data collection, a dietary food record was collected by phone call.

### Anthropometric Assessments

Measured anthropometric characteristics include body weight, body height, waist (WC), and hip circumference (HC) as proposed by Lohman ^29^. Body mass index (BMI) was calculated as the ratio between the body weight and the height squared (kg/m^2^). Interpretation of these results followed the international classification proposed by the World Health Organization ^30^.

### Bioelectrical impedance analysis

Body composition was assessed by using the bioelectrical impedance multiple-frequency (BIS) analysis (Body Composition Monitor – Fresenius Medical Care®), with different frequencies (5 to 1,000 kHz). The BIS analysis provided data regarding fat mass (FM), fat-free mass (FFM), phase angle (PhA), total body water (TBW), extracellular water (EW) and intracellular water (IW). It was calculated the ratio between EW and TBW as well. For the PhA, it was considered as worse values < 5.6º ^8^, and for the ratio between EW and TBW the overhydrated was considered as ECW/TBW ≥0.4 ^31^.

### Handgrip Strength

Handgrip strength (HGS) was assessed by the CharderMG4800 dynamometer. Participants were asked to sit comfortably with their shoulder adducted and forearm neutrally rotated, elbow flexed to 90°, and forearm and wrist in a neutral position using the dominant hand ^32^ or contralateral side to mastectomy, in the adjuvant cases, and lymphedema (BC group). The highest value of the three tests was used for the analysis ^33^. The interpretation of muscle weakness followed the classification proposed by a Brazilian cohort ^34^, in which values below < 16kg were classified as weakness. It was considered as “yes” for the weakness group participants whose HGS values were below the cutoff.

### Dietary data collection

The collection of dietary data occurred through a 24-hour food record for the study. It was collected 2 dietary records: the first one was collected on the day of the study visit and the second was collected after 3 weeks. The specific time frame was from the time the participant awoke in the morning until the time they slept at night. For this method it was used the methodology of the triple-pass 24-hour recall according to Nightingale et al ^35^, to improve the accuracy for quantification of the recall. The results obtained by the recall were inserted in the brazilian nutritional software Diet Box® to calculate the total amount of ingested energy and macronutrients. This software uses the Brazilian table of food composition in the assessment.

Reported values were analyzed by the Multiple Source Method (MSM) to estimate the usual intake distribution for daily-consumed nutrients. The MSM is a statistical method proposed in Europe by a German team [43] which accessible is through an open source online platform. By the probability of consumption and the amount consumed and regressions models, it corrects the within-person variance of the food intake results obtained by the record and yet it generates the usual intake for each participant [43]. Prior studies have shown that the MSM is an useful tool that provides usual nutrient and food intake estimates [44,45], thus, in order to improve the accuracy of the food consumption collected data, the MSM was applied. For the protein requirements and adequacy it was used for breast cancer patients the recommendation of 1.2 g/kg, as proposed by ESPEN guidelines ^36^. For the fiber requirements and adequacy, it was used for adult female recommendations being 25 g/d, according to a review with definitions and regulations for dietary fiber based on official recommendations by dietary reference intakes (DRIs) ^37^.

### Blood biochemical analysis

For the blood biochemical analysis it was asked to all groups, to fast for 12 hours previously. During the study visit at the hospital a nurse collected a 9ml tube of peripheral blood for the BC group and a researcher nurse collected it for MC and HC groups. This sample was processed in the nutrition and metabolism laboratory. The peripheral blood was collected, and serum was used for the following analysis: Albumin (AL); Total protein (TP); C-reactive Protein (CRP); fasting glucose (FG); Triglycerides (TG); High-density lipoprotein (HDL); total cholesterol levels (CT). For the low-density lipoprotein (LDL) it was used the Friedwald equation ^38^.

### Nutritional Risk Index (NRI)

The nutritional risk index was proposed in 1988 ^39^ in order to assess the nutritional status of participants through albumin levels. In 2005, this index was modified ^40^, introducing the ideal body weight into the formula. The NRI was calculated following the equation:

NRI = (1.519 × serum albumin, g/dL) + {41.7 × present weight (kg)/ideal body weight(kg)} The ideal body weight was calculated using the Lorentz formula for females ^41^:

Ideal weight = (height − 100) − ((height − 150)/2). In those cases that body weight was over than ideal weight, the fraction present weight (kg)/ideal body weight(kg) was adopted as 1 ^40^. Risk stratification the NRI for malnutrition was classified as: normal risk (≥100); mild risk (97.5 ≤ NRI<100); moderate risk (83.5≤ NRI <97.5); severe risk (NRI <83.5) ^40,42^. It was considered as “no” for patients with nutritional risk group, participants whose NRI values were below the cutoff (<100).

### Visceral Adiposity Index, Lipid Accumulation Product Index and Triglyceride Glucose Index

Metabolic disorders, insulin resistance, visceral fat dysfunction and lipid over accumulation were used to assess cardiovascular risk by using the triglyceride glucose index (TyG), visceral adiposity index (VAI), and lipid accumulation product index (LAP). TyG was calculated as described by Simental-Mendia et al (2008), according to the formula: TyG index = Ln (Natural logarithm) [(TG(mg/dL) × FG(mg/dL)/2] ^43^. VAI was calculated according to the formula for women: VAI = (WC(cm)/(36,58+(BMI *1.89) *(TG/0.81) *(1.52/HDL) ^44^, and LAP was calculated according to the formula for women LAP= [waist (cm)−58] × TG concentration (mmol/l) ^45^. For TyG index was considered as cutoff for metabolic syndrome and insulin resistance values >8.45 for females ^46^. VAI index classification considered as being “metabolically healthy” was defined as VAI <1.59, and “metabolically unhealthy” as VAI ≥1.59^47^. For LAP index classification it was considered LAP >30.40 as metabolically unhealthy ^48^.

### Blood pressure

The blood pressure (BP) was evaluated using automated cuff, the Omron device (HEM-7200) from the Omron 7000 line. Two measures were taken 60 seconds apart and repeat until both measures are within 6 mmHC for both systolic and diastolic.

### Statistical Analysis

The sample size calculation was performed using G*Power software version 3.1.9.4, taking into consideration the effect of chemotherapy on lipids status ^49^. The effect size of 0.575 showed that with a significance level of 95% and statistical power of 80%, using a Student t-test for paired data with a 2-sided significance level of .05. The minimum number of participants required was 29. Characteristics were summarized with the use of descriptive statistics such as mean, standard deviation (SD), median, and percentage. Shapiro-Wilk test was used to verify the distribution of continuous variables. Paired t-tests to compare the BC group to matched MC group, and two-tailed two-sample t-tests were used to compare BC group to HC group as well as MC females to HC females to analyze whether there was a statistically significant difference among the mean values of the variables of interest and to compare the differences among the groups. The analysis was run twice: the first test was considered the entire data, and in the second the outliers were removed. The results were the same for both, therefore the outliers did not influence the results reported. A level of significance was set at 0.05, and SAS Studio on SAS Institute Inc. 2015. SAS/IML® 14.1 User’s Guide was used for all data analysis.

## Results

Regarding the BC group, 36 females were included, 67% were Stage II, 28% were Stage III, and there were 5.5% at Stage I. The mean age was 45 years old (range, 26 – 64 years old), and the majority of women was younger than 50 years (69.5%). The prescibed protocol of treatment was the combination among Doxorubicin, Cyclophosphamide, and Docetaxel (AC-T). Clinic characteristics of the BC group are shown in table 2.

**Table 2:** Sample clinic characteristics.

| Variables | N | % |
| --- | --- | --- |
| Cancer stage |  |  |
| I | 2 | 5.50% |
| II | 24 | 67% |
| III | 10 | 28% |
| **Chemotherapy** |  |  |
| Neoadjuvant | 25 | 69% |
| Adjuvant | 11 | 31% |
| **Expected cycle numbers** |  |  |
| 4* | 1 | 3% |
| 8 | 35 | 97% |
| **Treatment Protocol** |  |  |
| AC-T | 36 | 100% |

\* One participant received a shorter protocol of chemotherapy. Caption: ACT – Cyclophosphamide, Doxorubicin and Docetaxel

There were no differences between the breast cancer patients and matched and health control in terms of age (45.3 years, 44.8 years, and 41.4 years respectively, P>0.05), and FFM (34.2 kg, 36.1 kg and 35.2 kg years respectively, P>0.05). Regarding of weight, BMI, WC and FM, there were also no differences between BC and MC (P>0.05). According to BMI classification and FM results, it was observed a high prevalence of overweight and obesity in the BC group as well as in the MC, and for both measurements (BMI and FM) there were significant differences when both BC and MC were compared to CH (P<0.05). BC females also differed from the MC and HC in terms of PhA and EX/TBW results, in which BC had the lowest values for PhA (5.3). The non-malignancy groups (MC and HC) presented better values of HCS, NRI and BP as well. Table 3 present the complete data of the anthropometric, body composition, nutritional risk, HGS, and blood pressure among the groups.

**Table 3:** Sample characteristics.

| Variable | BC<br>n=36) |  | MC<br>(n=36) |  | HC (n=16) |  | Significance |  |  |
| --- | --- | --- | --- | --- | --- | --- | --- | --- | --- |
|  | Mean | SD | Mean | SD | Mean |  | BC vs<br>MC | BC vs<br>HC | MC vs<br>HC |
| Age (y) | 45.3 | 8.9 | 44.8 | 9.0 | 41.4 | 9.9 | 0.23 | 0.18 | 0.23 |
| Height (cm) | 159.3 | 7.7 | 162.9 | 5.8 | 161.5 | 7.8 | **0.03** | 0.37 | 0.47 |
| **Weight (kg)** | 74.0 | 13.8 | 76.6 | 15.3 | 59.8 | 8.2 | 0.1 | **0.03** | **0.01** |
| **BMI (kg/m<sup>2</sup>)** | 29.0 | 4.9 | 28.6 | 4.7 | 22.2 | 2.7 | 0.11 | **0.001** | **0.001** |
| **WC (cm)** | 95.4 | 10.5 | 94.4 | 11.8 | 82.6 | 8.1 | 0.56 | **0.001** | **0.007** |
| **FFM (KG)** | 34.2 | 9.5 | 36.1 | 6.2 | 35.2 | 7.3 | 0.28 | 0.7 | 0.65 |
| **FM (KG)** | 28.1 | 9.6 | 30.1 | 11.3 | 18.9 | 4.4 | 0.25 | **0.02** | **<0.001** |
| **PhA** | 5.3 | 0.8 | 6.4 | 0.8 | 6.3 | 0.9 | **<0.001** | **<0.001** | 0.69 |
| **TBW** | 33.3 | 7.1 | 33.5 | 4.8 | 30.3 | 7.5 | 0.83 | 0.18 | 0.06 |
| **EW** | 15.8 | 3.5 | 14.7 | 2.3 | 13.3 | 3.4 | **0.03** | **0.02** | 0.08 |
| **IW** | 17.5 | 3.9 | 18.5 | 2.7 | 17.0 | 3.7 | 0.17 | 0.7 | 0.12 |
| **EW/TBW** | 0.5 | 0.0 | 0.4 | 0.0 | 0.4 | 0 | **<0.001** | **<0.001** | **0.04** |
| **BPsys [mmHC]** | 122.1 | 16.1 | 115.5 | 13.8 | 111.3 | 10.3 | 0.06 | **0.02** | 0.29 |
| **BPdia [mmHC]** | 80.7 | 11.6 | 74.3 | 9.0 | 71.1 | 10.5 | **0.02** | **0.008** | 0.28 |
| **HGS (kg)** | 22.5 | 5.7 | 27.1 | 6.3 | 24.4 | 5.1 | **<0.001** | 0.19 | 0.39 |
| **NRI** | 93.4 | 9.5 | 101.4 | 7.0 | 98.0 | 7.4 | **<0.001** | 0.14 | 0.11 |

BC: Breast cancer group, MC: Matched control group, HC: healthy control group, WC: waist circumference, BMI: Body mass index, HGS: Handgrip strength, FFM: Fat-free mass, FM: Fat mass, PhA: Phase angle. TBW: Total body water. EW: Extracellular water. IW: Intracellular water. EX/TW: The ratio between extracellular water and total water BP sys: Systolic blood pressure. BP dia: Diastolic blood pressure. NRI: Nutritional risk index \* The mean difference is significant at a level of 0.05.

Considering the nutritional markers tools, BC females had the highest prevalence of inadequacy and critical values for all measurements (PhA; HGS and NRI) when compared with matched and healthy control. MC and HC had no difference in any of those markers. Figure 1 shows those comparisons.

**Fig 1a:**
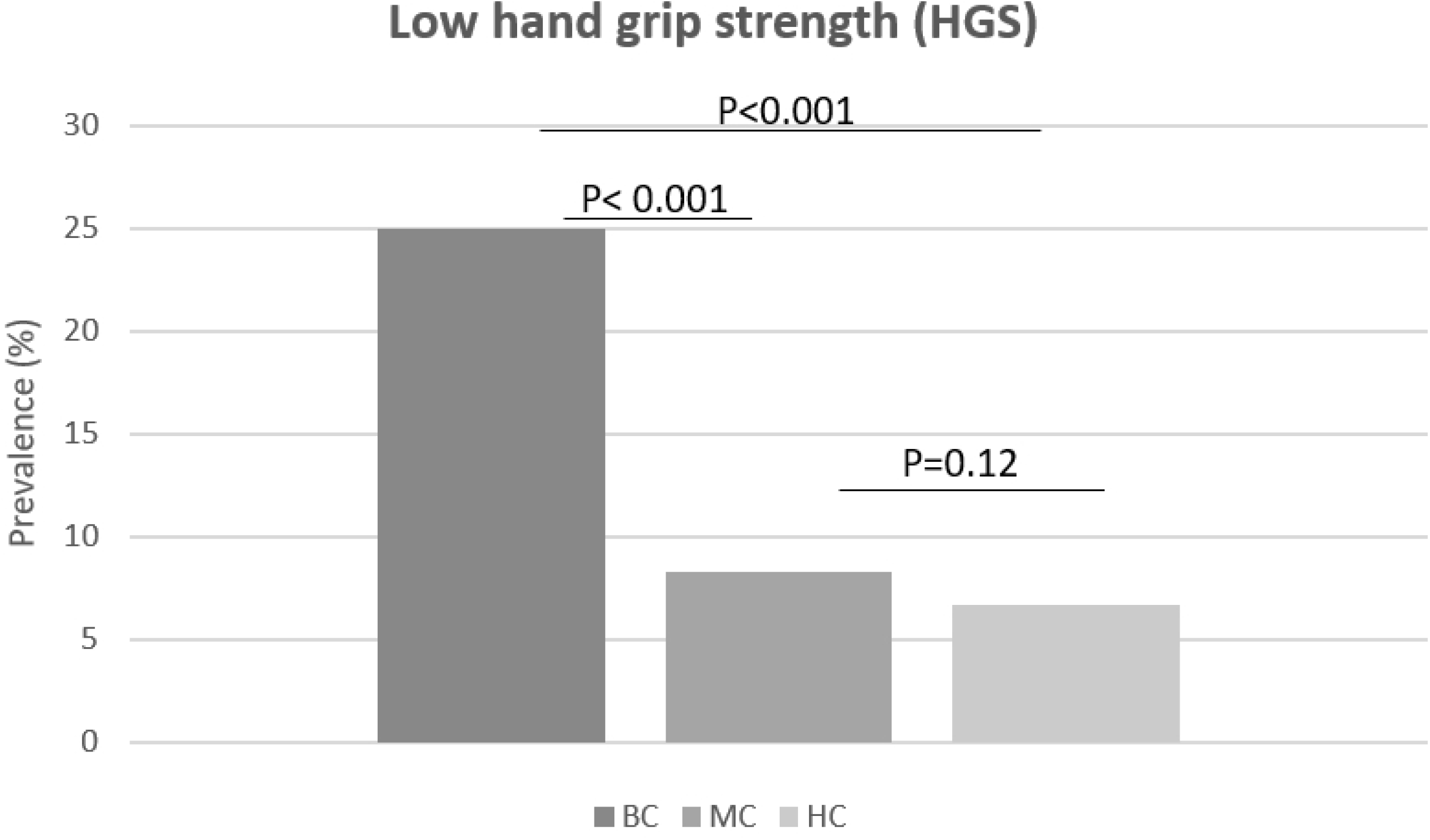
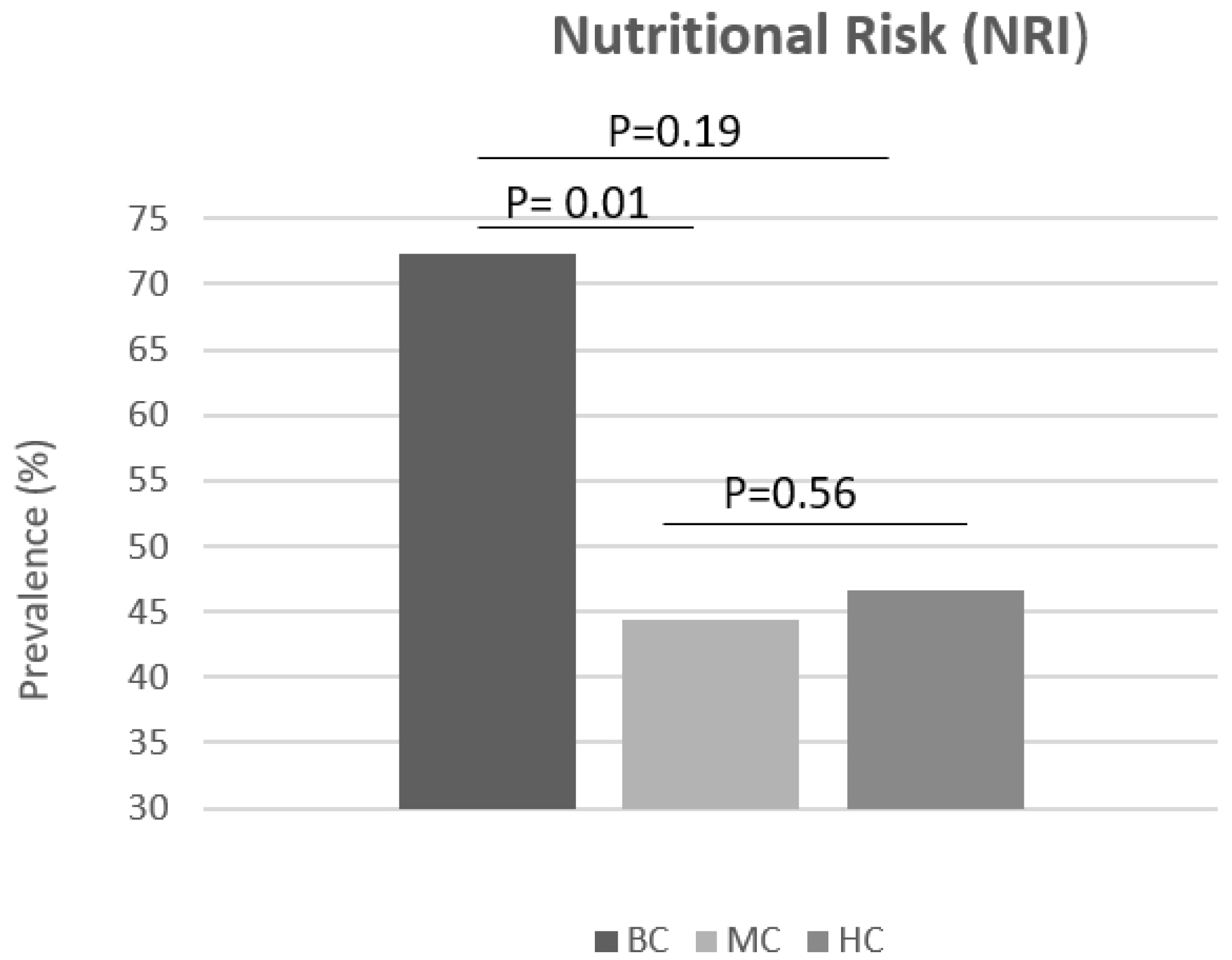
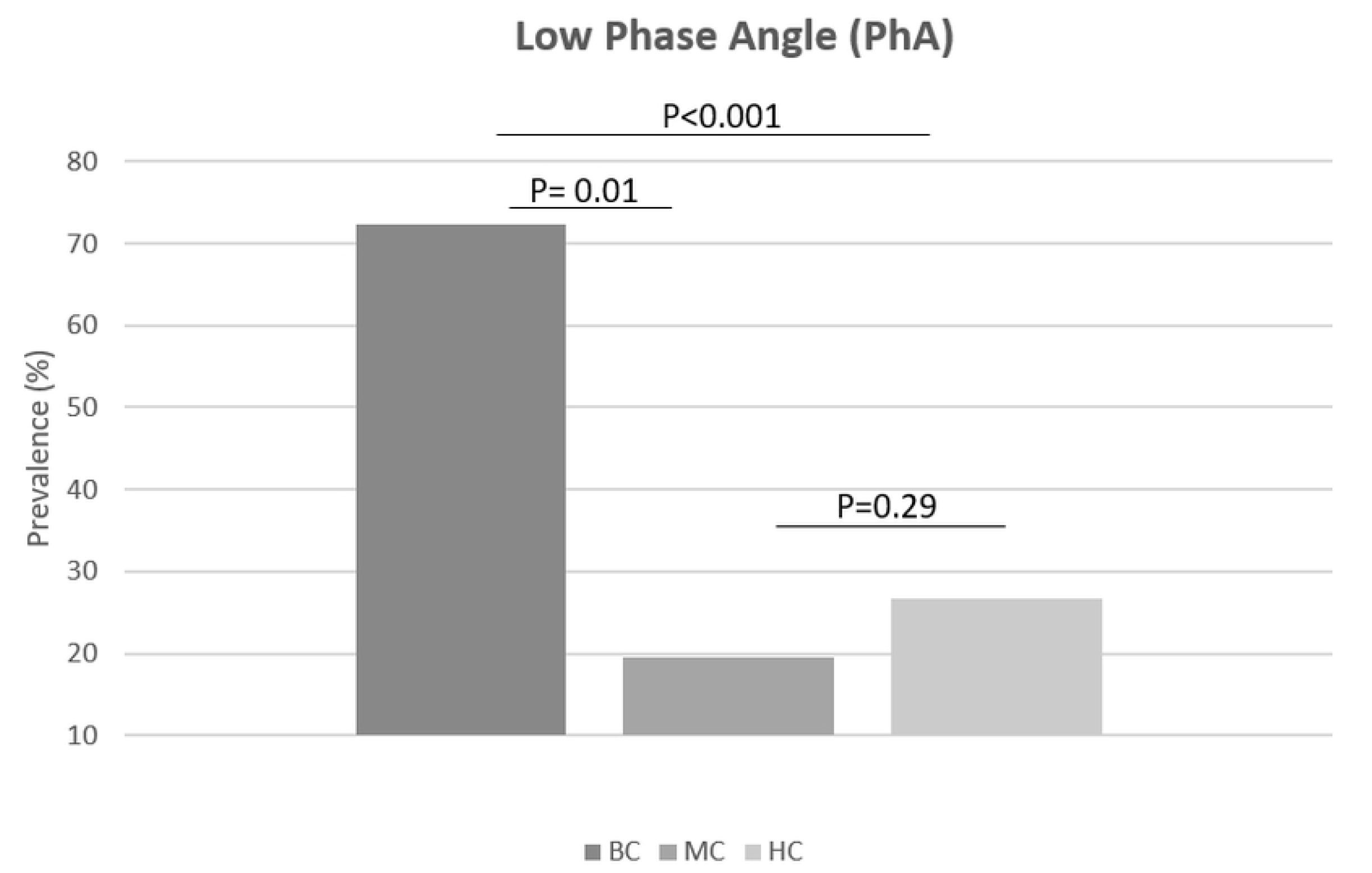
Prevalence of low handgrip strength; Fig 1b: Prevalence of nutritional risk; Fig 1c: Prevalence of low phase angle values. Captions: BC: Breast cancer patients; MC: Matched control group; HC: Healthy control group. The mean difference is significant at a level of 0.05.

Regarding the food consumption of the groups, daily caloric and carbohydrate intake were higher in the BC group (1744 kcal and 245.2 g respectively) and differ in statistically significance from MC for both values. The range of protein intake/kg for BC group was 0.3 g/kg – 1.9 g/kg, and only 41.6% of the patient (N=15) achieved the minimal recommendation from Espen guidelines. Interesting, BC females also were the group with the highest intake of fiber, being statically significant when compared to the matched group (P=0.005). Additionally, the other macronutrient distribution did not differ between any participant groups. Table 4 shows the complete food intake results and their comparisons among the groups.

**Table 4:** Food intake results among the groups.

| Variable | BC n=36) |  | MC (n=36) |  | HC (n=16) |  | Significance |  |  |
| --- | --- | --- | --- | --- | --- | --- | --- | --- | --- |
|  | Mean | SD | Mean | SD | Mean | SD | BC vs MC | BC vs HC | MC vs HC |
| **Energy (kcal)** | 1744.3 | 559.1 | 1363.8 | 572.6 | 1590.2 | 498.0 | **0.004** | 0.35 | 0.19 |
| **CHO (g)** | 245.2 | 93.0 | 177.9 | 729.2 | 177.8 | 93.2 | **0.001** | **0.02** | 0.99 |
| **Protein (g)** | 77.9 | 24.0 | 72.7 | 39.1 | 78.5 | 25.9 | 0.5 | 0.93 | 0.59 |
| **Protein g/kg** | 1.1 | 0.0 | 1.0 | 0.6 | 1.3 | 0.3 | 0.37 | 0.1 | 0.08 |
| **Total fat (g)** | 50.7 | 21.0 | 47.3 | 22.3 | 48.5 | 23.1 | 0.51 | 0.74 | 0.86 |
| **Col (g)** | 281.5 | 141.7 | 255.3 | 220.0 | 262.1 | 184.9 | 0.99 | 0.69 | 0.83 |
| **Fiber (g)** | 15.9 | 7.5 | 10.4 | 6.9 | 13.3 | 7.5 | **0.005** | 0.26 | 0.19 |

Captions: Breast cancer group; MC: Matched control group; HC: healthy control group; CHO: Carbohydrate; Protein g/kg: it was considered the ratio between the total amount of protein and body weight for each participant. Col: Cholesterol. \* The mean difference is significant at a level of 0.05.

Concerning about biochemical results, and overall, BC group had the worst value among all analyses, and it was significantly different from the matched group in regarding of FG, TG, HDL, TC, TP, and albumin results (P<0.05). MC and HC did not differ in any biochemical parameters. Table 5 presents the complete data of biochemical test and their variance among the group results.

**Table 5:**
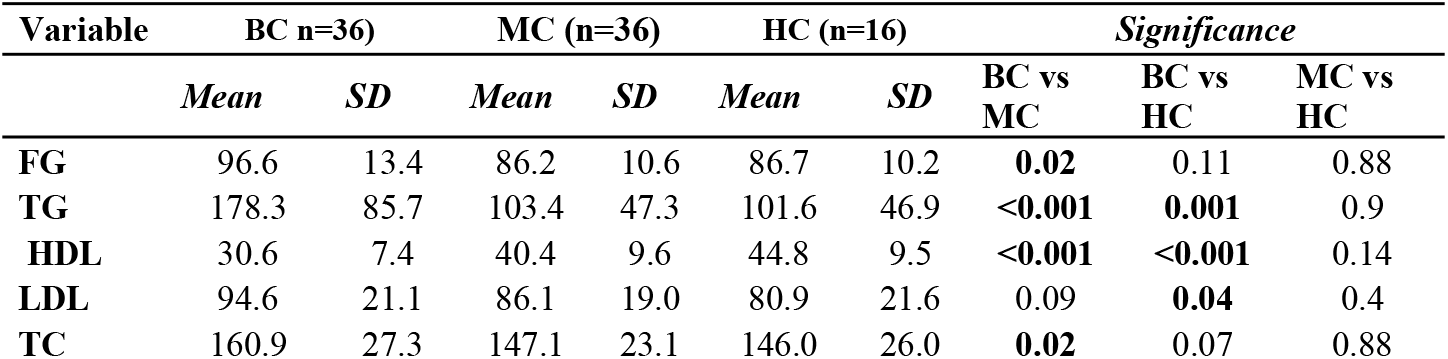
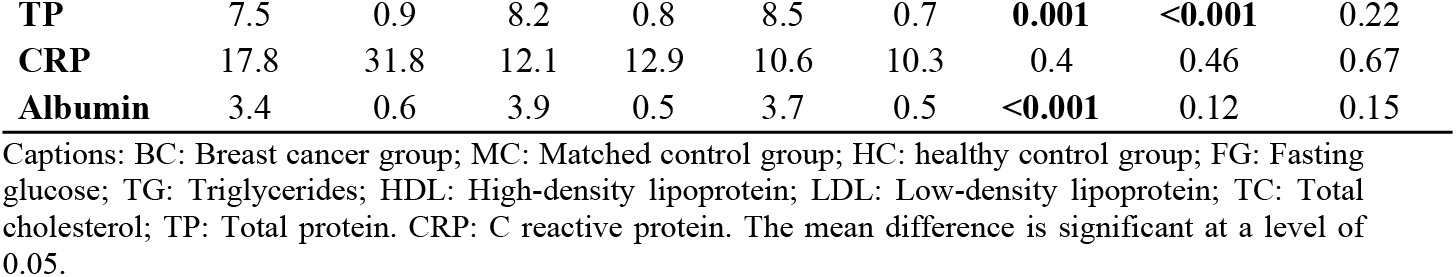
Biochemical test and their variance between the groups.

As expected, following the metabolic differences observed in table 5, BC group patients had worse values of all adiposity markers, for both adiposity index (VAI and LAP), and metabolic and cardiovascular risks measured by TyG index. For the three indices, the mean value of BC group was statistically higher (P<0.05). Furthermore, it was not found difference regarding MC and HC groups for those evaluations. Figure 2 present the distribution of VAI, LAP and TyG among the groups.

**Fig 2a:**
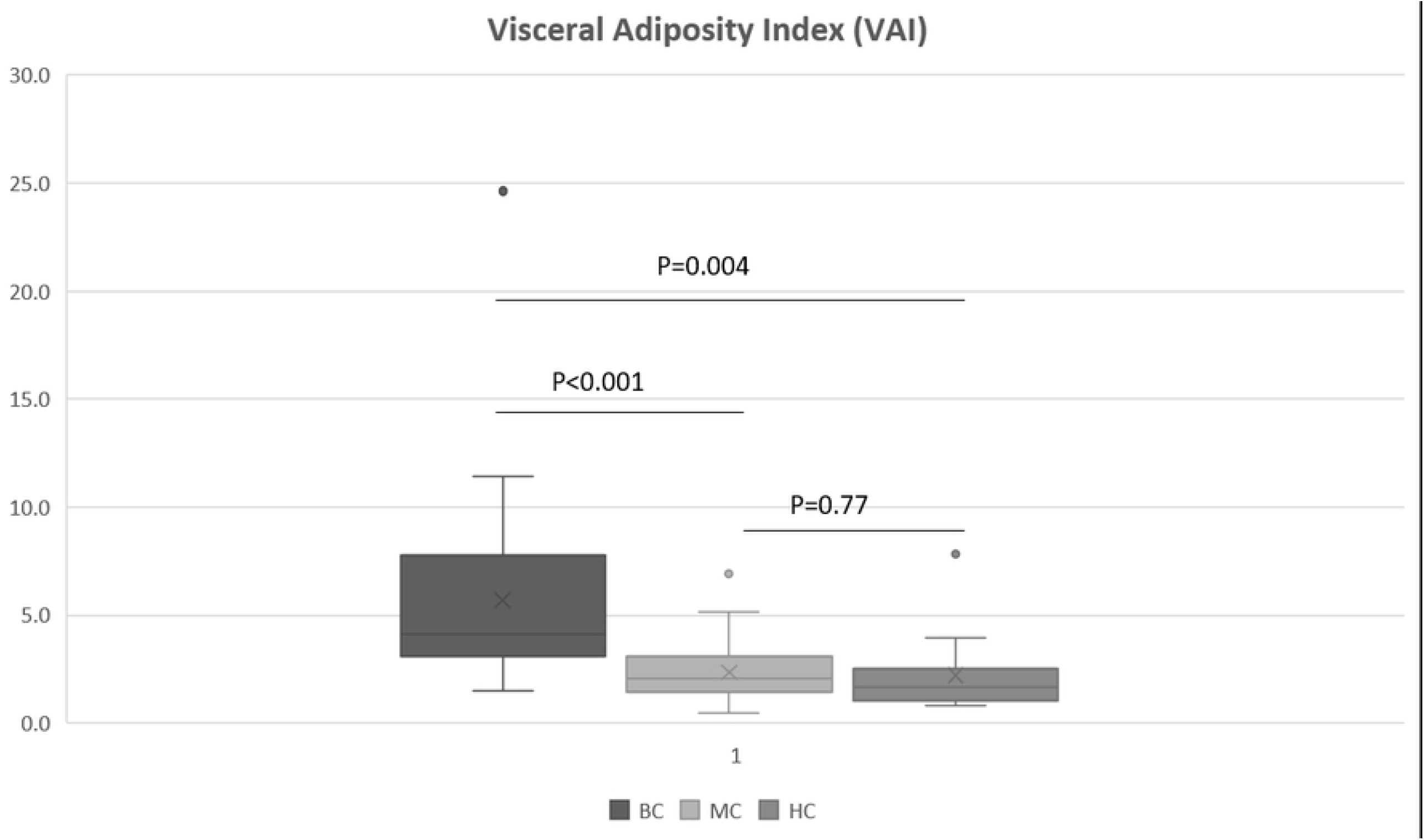
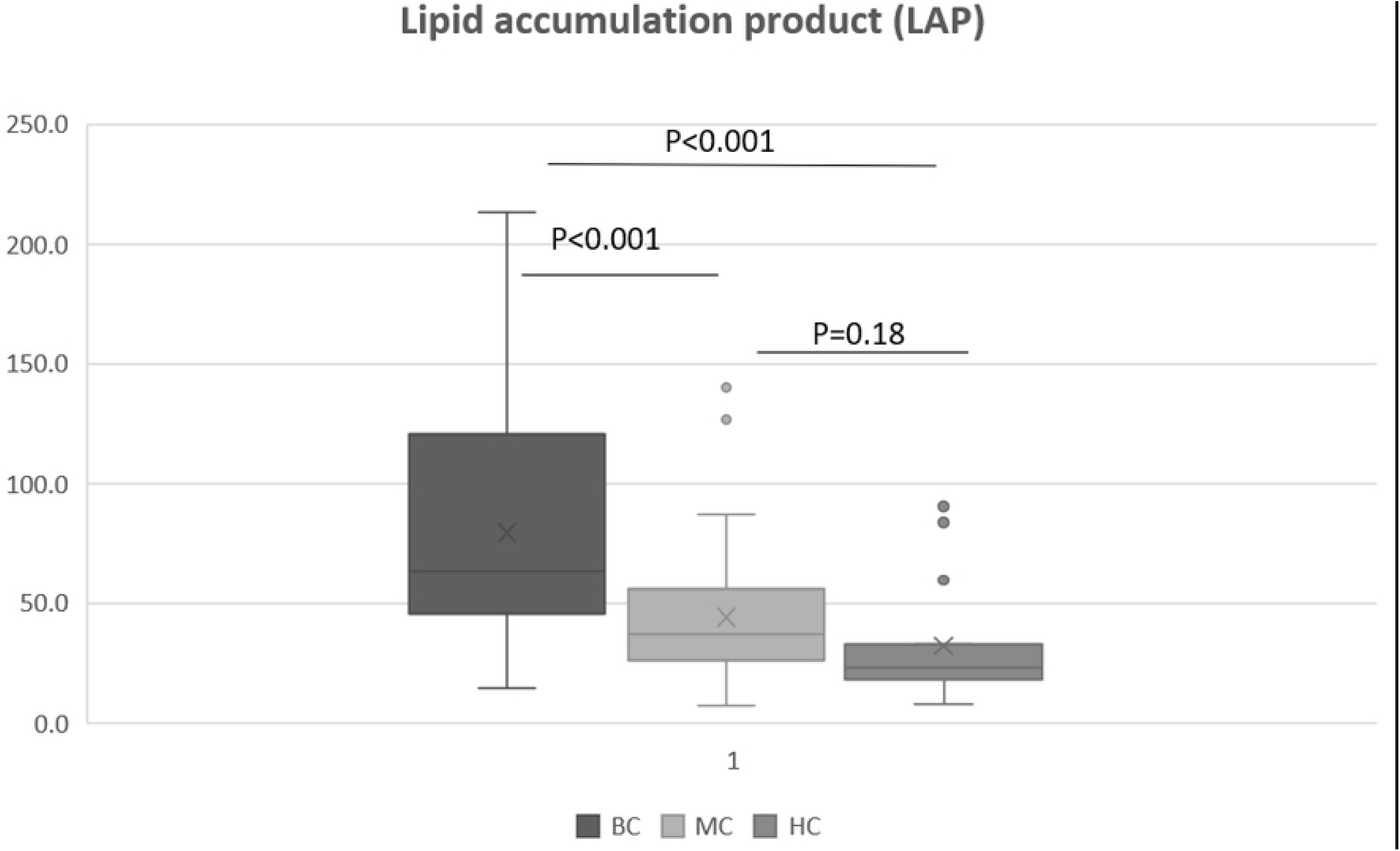
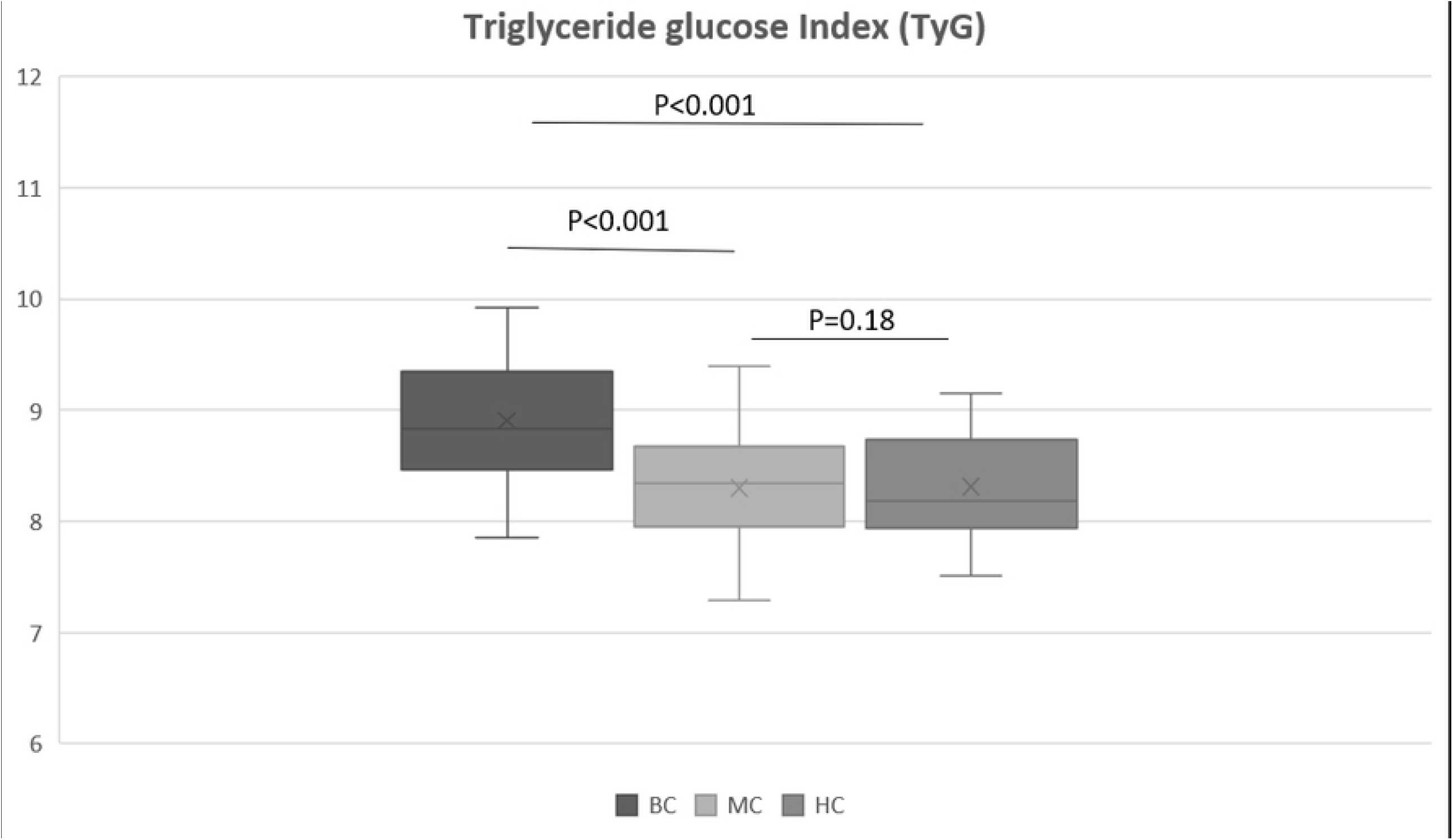
Distribution of visceral fat index; Fig 2b: Distribution of lipid accumulation index; Fig 2c: Distribution of triglyceride glucose index. Captions: BC: Breast cancer patients; MC: Matched control group; HG: Healthy control group. The mean difference is significant at a level of 0.05

## Discussion

To our knowledge, only a few articles aimed to make similar comparisons. A meta-analysis conducted by Hernandez et al (2014) identified 22 studies in which compared breast cancer patients with a control group ^50^. However, only 2 studies included dietetic data besides the metabolic factors comparisons ^51,52^, and we were the only study conducted with Brazilian population in which included both body composition and nutrition status parameters. Breast cancer patients on average are presented with a high prevalence of abdominal obesity, high body weight, BMI and fat mass, and consequently the matched control as well. Both groups differed from the healthy control group in all of those obesity indicators.

The results of our study indicate that, despite similar age, BMI, waist and hip circumferences fat-free mass and fat mass (not statically significant), it was possible to verify impairments in aspects of nutritional status markers among BC and matched control group, but not between the MC and HC groups. BC group presented the lowest values of PhA, and the highest prevalence of low HGS, nutritional risk by NRI and overhydration by EW/TBW. All those parameters are considered indicators of poor nutritional status ^8,53–55^. Regarding PhA, the BC group had values lower than the cutoff proposed by Gupta et al (2008) in which values below 5.6 are considered a sign of poor prognosis for breast cancer patients ^8^. PhA values for both healthy controls did not differ as well as NRI and HGS values. Besides nutritional status, breast cancer patients on average presented poor indicators of metabolic health, with the highest levels of BP, FG, all lipids’ markers, CRP, and the lowest level of albumin. Despite significant differences in body weight, WC, and fat mass levels, MC and HC did not differ in any biochemical parameters. This result is concordant with previous research that has already reported differences in glucose metabolism and metabolic syndrome prevalence among breast cancer patients ^51^, as well as alterations in insulin homeostasis when compared to the control group ^56^.

Obesity is a condition that is frequently associated with abnormalities in lipid metabolism ^57^, however, in this study, we did not find lipids impairments in the MC group, only among the cancer patients. In addition to body composition, other components can contribute to lipids alterations, as the own tumor, in which lipid metabolism changes influence proliferation and dissemination of cancer cells ^58^, and chemotherapy itself has the potential to promotes modifications in serum lipids ^59^. Thus, those components may explain the reason of presence of lipids alterations only in BC group. Moreover, poor metabolic indicators contribute to increasing the risk of various conditions such as atherosclerosis, and other cardiovascular diseases ^24^, and according to Bell et al (2014) metabolic syndrome components can increase risk of death by 3 times ^51^.

In order to verify and compare cardiovascular risk, we included in this study adiposity and lipid accumulation indices. Several studies have already shown the VAI, LAP and TyG indices as simple and good markers of cardiovascular outcomes and as screen tools for cardiovascular disease risk ^60–65^. Kouli et al (2017), during a 10-year follow-up of a cohort of 3,042 adults, found that VAI was independently associated with an elevated risk of CVD in 10 years ^60^. Furthermore, considering the TyG index, a study with a Brazilian population found superior performance compared to the HOMA method for estimation of insulin resistance ^66^. In this study, as expected according to the discrepancies of biochemical blood results among the groups, the BC had the worst value of VAI, LAP and TyG, indicating a visceral fat dysfunction, therefore, high cardiovascular disease risk. Additionally, despite the difference in body composition, it was not found any difference of those indices among the MC and HC.

We attempted to identify possible reasons for the difference in the metabolic and nutritional markers between patients and MC females by measuring food intake as body composition did not influence those results (MC group presented healthier results than BC). There have been many studies performed outlining the role of diet and disturb on glucose and lipids metabolism. A review conducted by Siôn A.P & Hodson L (2017) concluded that energy intake, independent of nutrient content, is a crucial regulator of hepatic lipid accumulation ^67^. Furthermore, CHO levels in diet are also responsible for metabolic profile alterations where high ingestion is associated with an increase in serum lipids ^68,69^. Concordantly, in this study, we confirmed breast cancer patients had the highest level of energy and carbohydrate intake, and it was statistically different from the matched group intakes and from HC regarding CHO consumption, however, considering caloric intakes, it did not differ from the healthy females. We hypothesized that along the reasons we did not find discrepancies in caloric ingestion among patients and the healthy control. There could be differences in the energy expenditure and level of physical activity, notably, and, as a limitation of our study, we did not evaluate those components.

Curiously, BC presented the highest level of fiber ingestion, despite being far from the 25 g/day recommendation, evidencing the diet inadequacies among the population overall. In addition to the dietary shortcomings observed, the patients had low protein intake/kg as well. Although it was not statistically significant when compared to the other groups (MC and HC), it is still clinically relevant, especially considering that changes in nutritional status markers have already been identified in this sample (PhA, NRI and EW/TBW) and low HGS. Besides, there is a potential for the development of sarcopenic obesity in those patients with an intake below 1.2g / kg ^70^.

Contrary to our hypotheses, we observed no differences in body composition (FM and FFM) between breast cancer patients and matched females, and also no differences in FFM among the 3 groups, though BC still had a higher nutritional risk. However, as we hypothesized, cancer patients demonstrated impairments in lipids, worst glucose levels, visceral fat dysfunction and consequently higher cardiovascular risk in which those females presented important unhealthy dietary patterns with higher carbohydrate and caloric intake and insufficient protein and fiber ingestion. Accordingly, our findings highlight the need for the implementation of a targeted dietetic approach to treat and mostly to prevent unfavorable metabolic and nutritional outcomes. Successfully, dietetic management has already shown to be an effective method to control and prevent metabolic impairments ^71–74^; it is particularly important in the breast cancer patient population where the metabolic risks are increased by the tumor and chemotherapy besides the diet and body composition unfavorable. Remarkably our study has already mentioned-limitations as not inclusion of energy expenditure and physical activity investigation and the sample size was relatively small. For further studies, the inclusion of those components and expanding the follow-up of these patients could better elucidate the mechanism involved in metabolic changes and whether these results are maintained in the long term or enhanced by hormone therapy.

Finally, this study has found that women undergoing breast cancer chemotherapy, after completion of the treatment, presented poor indicators of nutritional and metabolic health, such as PhA, NRI, EW/TBW HGS, dyslipidemia, and visceral fat dysfunction by adiposity indices when compared to a group of age- and BMI-matched non-malignant females. Body composition and age do not explain these differences. Furthermore, the dietetic investigation revealed a higher energy intake and carbohydrate and insufficient consumption of protein and fiber.

Considering the possibility of poor prognosis related to the nutritional markers, sarcopenic obesity or the subsequent threat of developing cardiovascular disease in survivorship, this study highlights the necessity for more effective lifestyle intervention as exercise and nutrition counseling during breast cancer treatment.

## Data Availability

All relevant data are within the manuscript.

## Declarations

### Authors’ contributions

The authors’ responsibilities were as follows – AAJJ: conceptualized the study; BRS, LAPC, TOG, and AAJJ: were responsible for the research design; BRS and LAPC: conducted the research and analyzed the data; BRS, MM, and AAJJ: wrote the paper and had primary responsibility for final content; and all authors: contributed to data interpretation and read and approved the final manuscript.

### Ethical standard

All human studies have been approved by the appropriate ethics committee and have, therefore, been performed in accordance with the ethical standards laid down in the 1964 Declaration of Helsinki and its later amendments.

All persons gave their informed consent prior to their inclusion in the study.

### Availability of data and material

All relevant data are within the paper

### Code availability

Not applicable

## Acknowledgments

We thank all of the research group on Nutrition and Breast Cancer of the University of São Paulo, especially the students who assisted in all phases of the study.

